# From Genes to Neurochemistry: Excitation and Inhibition Mechanisms of Sensory Differences in Autism

**DOI:** 10.64898/2026.07.14.26358047

**Authors:** Alice R. Thomson, Viola Hollestein, Martina Arenella, Helen Powell, Jason He, Beth Oakley, Eva Loth, Rosemary Holt, Jan K. Buitelaar, Laura Colomar, Natalie J. Forde, Thomas Bourgeron, Terje Falck-Ytter, Giorgia Bussu, Tobias Banaschweski, Pascal-M Aggensteiner, Richard Edden, Tony Charman, Charlotte Pretzsch, Declan Murphy, Tomoki Arichi, Nicolaas A. Puts

## Abstract

Sensory processing differences are a core feature of autism, affecting 60–95% of individuals, yet the associated neural mechanisms remain unclear. An excitation–inhibition (E/I) imbalance in brain circuits has been proposed, but in vivo evidence linking genetic variation in E/I pathways, regional neurochemistry, neural circuit function, and sensory behaviour has been lacking. Here we performed a multimodal investigation in 206 individuals (130 autistic), integrating gene-set polygenic scores for excitatory glutamatergic and inhibitory gamma-aminobutyric acid (GABA)-ergic pathways, magnetic resonance spectroscopy (MRS) measures of regional GABA and Glx (glutamate + glutamine) levels, vibrotactile psychophysical measures of tactile perception, and questionnaire measures of behavioural sensory reactivity. We found that glutamatergic polygenic scores predicted thalamic glutamate levels in neurotypical but not autistic individuals, suggesting altered genotype–neurochemistry coupling in autism. Thalamic Glx:GABA levels associated with tactile perception in both groups, but with opposing directions of effect, indicating that autistic and neurotypical individuals achieve similar perceptual outcomes with potentially differing thalamocortical circuit mechanisms. Within autistic individuals, tactile perceptual differences further related to behavioural sensory reactivity. Together, these findings suggest that autistic sensory processing potentially relies on distinct circuit mechanisms linking genetic variation, neurochemistry and perception. This work thus has important implications for how sensory differences are conceptualised, studied, and interpreted, and ultimately for how interventions and support are developed.

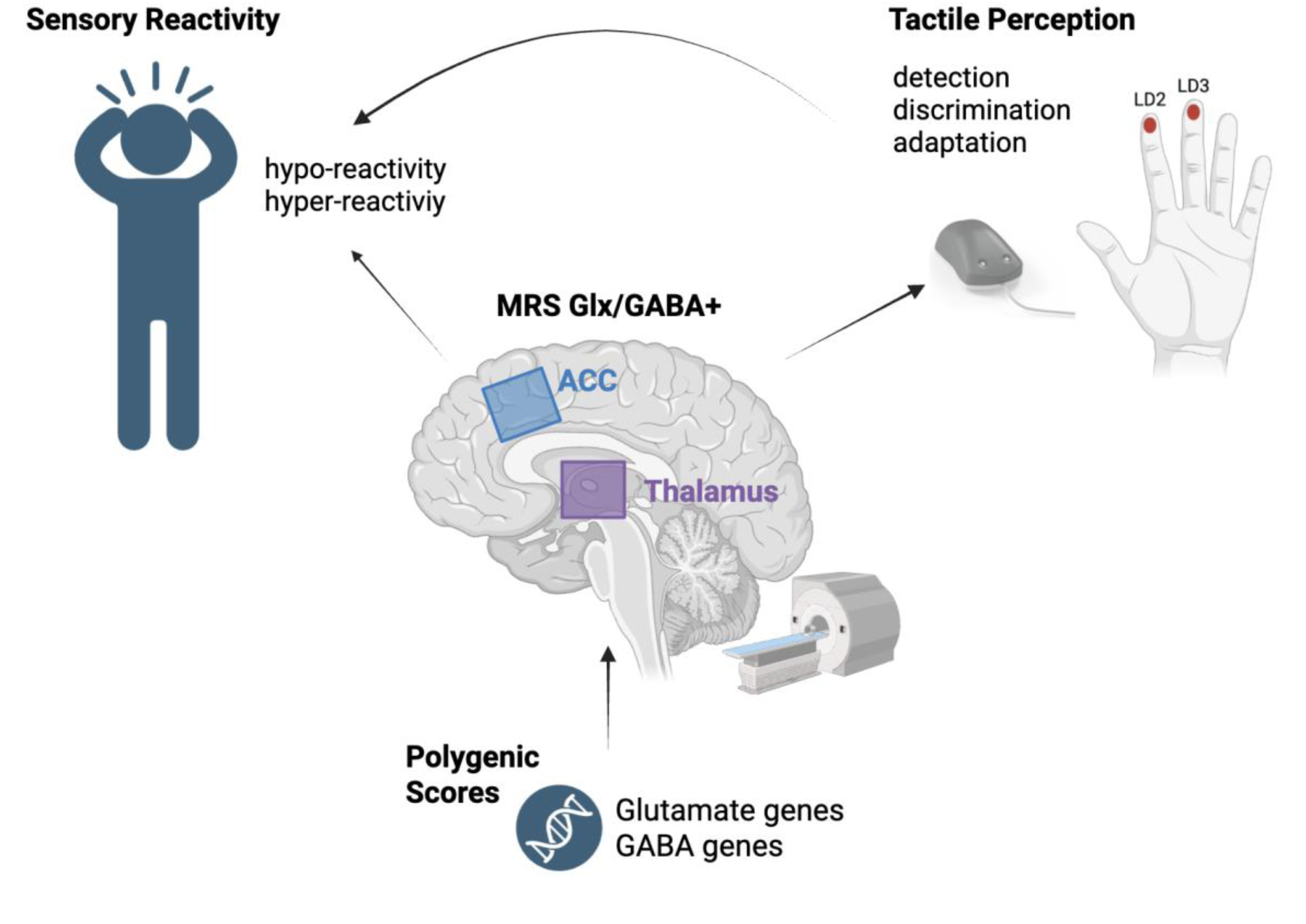

**Summary Figure:** Summary of our comprehensive investigation of the role of excitation and inhibition in sensory processing differences in autism by integrating GABA and glutamate gene set polygenic scores (PGS), MRS-measured thalamic and anterior cingulate cortex (ACC) Glx (glutamate + glutamine) and GABA+ (GABA + macromolecules) levels, vibrotactile psychophysical measures of tactile perception (tactile detection thresholds, discrimination thresholds and tactile adaptation etc.) and questionnaire-based measures of sensory reactivity (behavioural and emotional responses to sensory stimuli, including hyper-and hypo-reactivity).

## Introduction

Sensory processing differences are extremely common in autism, affecting 60–95% of individuals and spanning visual, auditory, tactile, taste and olfactory sensory domains (1,2). Examples of atypical sensory processing include sensory seeking behaviours (e.g., seeking bright lights or loud sounds), sensory avoidance (e.g., avoiding certain textures or sounds), and differences in sensory perception (3–8). Although sensory processing differences are among the earliest indicators of autism (observable as early as 7 months of age; (9–11), have been identified as a key research priority for the autistic community (12), and likely contribute to other core autistic traits (13,14), the neural mechanisms giving rise to sensory perceptual and behavioural differences *in vivo* remain poorly characterised.

A prominent theoretical framework proposes that autistic behaviours, including sensory differences, arise from an altered balance within neural systems between excitatory glutamatergic and inhibitory gamma-aminobutyric acid (GABA)-ergic neurotransmission (15). This excitatory/inhibitory (E/I) imbalance hypothesis is supported by an enrichment of autism-associated variants in excitatory and inhibitory synaptic gene pathways (16,17), as well as converging evidence from animal models and postmortem studies demonstrating alterations in excitatory and inhibitory synaptic receptor function and density (18,19). However, despite its widespread influence, the E/I imbalance hypothesis has rarely been tested across multiple levels of biological organisation in humans. In particular, it remains unclear how common genetic variation in E/I-related pathways translates into regional excitatory and inhibitory neurochemistry *in vivo* and ultimately into sensory experiences.

The touch perceptual domain is a particularly well-suited model sensory system for such a multi-level mechanistic investigation. Tactile perception (or low-level sensitivity) can be objectively measured using validated vibrotactile psychophysical paradigms, which probe different aspects of perception including tactile detection, adaptation and amplitude discrimination thresholds within the flutter range of touch (< 50Hz ; 14,15). Studies using these tasks report reproducible increases in low-level tactile perception thresholds in autistic individuals, suggesting potential differences in tactile stimulus encoding and/or processing. Perceptual paradigms can be combined with questionnaire-based measures of sensory reactivity, including affective reactivity (short-term emotional responses to sensory stimuli) and behavioural responsivity (longer-term behavioural responses such as avoidance; 16), enabling investigation across multiple levels of sensory processing.

The different vibrotactile perception paradigms also serve as proxy measures of specific cortical and subcortical inhibition mechanisms, thus allowing researchers to link perceptual processing to various underlying neural circuits (5,21,22). For example, vibrotactile amplitude discrimination tasks are thought to engage lateral inhibition within the primary somatosensory cortex (S1). This form of inhibition is mediated by gamma-aminobutyric acid (GABA)-ergic interneurons which bridge neighbouring cortical mini-columns (whose receptive fields encode distinct stimuli) and scale their activity relative to the surrounding neural population. These interactions enhance contrast between adjacent receptive fields, sharpening spatial resolution and facilitating the discrimination of simultaneous tactile inputs to neighbouring digits (23–29). The observed tactile discrimination differences in autism thus implicate altered inhibitory function, consistent with the E/I imbalance hypothesis.

Genetic findings also implicate GABAergic function in tactile perception, with single nucleotide polymorphisms (SNPs) in GABA_A_ receptor subunit genes (GABRB3 in particular), associated with both parent-reported reactivity and tactile detection thresholds to Von Frey fibres in NT children (30). In fact many of the gene implicated in autism pay essential roles in early neurodevelopmental processes (synaptogenesis, synaptic adhesion and stabilities), and are also associated with sensory sensitivity in animal models (31,32). Genome-wide association studies have identified approximately 100 autism loci (17), enabling the construction of polygenic risk scores (PRS) to estimate relative genetic liability. It is possible to summarise common genetic liability limited to GABAergic and glutamatergic gene sets (33), faciliating specific exploration of the role of autism assoicated genetic variation in E/I systems. This however has not yet been explored in the context of sensory phenotypes.

Further evidence supporting the role of an E/I imbalance in sensory differences in autism comes from studies using magnetic resonance spectroscopy (MRS), which enables the non-invasive quantification of regional brain GABA and glutamate (often quantified as glutamate + glutamine (Glx)). Glx and GABA can be measured as proxies of regional E/I activity, broadly reflecting excitatory and inhibitory tone; the extent to which a region can exert excitation or inhibition, representing tonic, metabolite and potentially phasic E/I activity (34). Prior studies report associations between MRS measurements of these neurochemicals, tactile perception, and broader questionnaire measures of sensory reactivity differences in autism (21,35,36).

Importantly however, these previous studies tend to focus on a single vibrotactile psychophysical measure, a single sensory questionnaire, or a single MRS proxy of excitation or inhibition in isolation. This approach overlooks the likely interactive nature of sensory difficulties (including sensory perception and reactivity differences) and E/I dynamics. GABA and glutamate are metabolically and functionally interdependent, and their combined activity shape’s regional neural function (37,38). Previous studies measuring GABA or Glx in a single brain region, without comparison to other regions, also lack a control to determine whether observed associations are region-specific. Furthermore, although autism-associated genes are enriched in GABAergic and glutamatergic pathways (16,17), it is unknown whether common genetic liability explains individual differences in *in vivo* measures of excitatory and inhibitory neurochemistry. The field thus lacks integrated investigations linking genetic variation, neurochemistry, perceptual processing and sensory behaviour within the same cohort, leaving the mechanistic pathway from genetic variation to sensory experience largely uncharacterised.

*Here*, we address these gaps by studying individuals from one of the largest autism cohorts to date (the Longitudinal European Autism Project (n = 206; 130 autistic; 13.5–37.5 years; LEAP - (39)). We use a multimodal approach that integrates individual polygenic scores from GABA and glutamate gene sets, MRS measures of GABA and Glx from the thalamus and anterior cingulate cortex (ACC), multiple vibrotactile psychophysical assessments of low-level tactile perception, and questionnaire-based measures of behavioural sensory reactivity. The MRS thalamus voxel was chosen based on its known role in the relay of sensory information from the periphery to the cortex (40). The ACC was selected because of its central role in higher-order sensory processing through its structural and functional connections with the prefrontal cortex, motor regions, and insula (41). In particular, the ACC is thought to encode the affective qualities of sensory stimuli, such as pleasantness (42,43), with altered ACC activity previously associated with tactile hypersensitivity (Lee et al., 2024). Specifically, we examine how common genetic variation shapes individual E/I neurochemical development in these regions, how regional E/I neurochemistry relates to vibrotactile perception, and then how perceptual differences contribute to sensory reactivity behaviours in autism.

## Materials and Methods

### Participants

Data were acquired as part of the LEAP Wave 3 (2020–2024), within the AIMS-2-TRIALS research programme (https://www.aims-2-trials.eu/; 30,33). Our sample consisted of 206 participants (130 autistic, 76 NT; 13.5 – 37.5 years). Data were acquired from 4 sites (on 4 different 3 Telsa MRI scanners; Table S1). Demographics are presented in Table 1 for the data included in this study. All participants had an intelligence quotient (IQ) > 75 (Wechsler Abbreviated Scale of Intelligence). For a detailed overview of study inclusion and exclusion criteria and overall protocol please refer to (39,47) and the supplementary materials. The research protocol was approved by the local research medical-ethics committees (39), all data were collected after written informed consent from the volunteers or a legal guardian.

**Table 1.** Participant demographics & voxel count per site. Note IQR = interquartile range.

| Site | N | Age (median (IQR)) | Sex (male (female)) | MRS voxel count (ACC:thalamus) |
| --- | --- | --- | --- | --- |
| Site 1 - NT | 13 | 23.0 (9.92) | 7 (6) | 8:13 |
| Site 1 - Autism | 45 | 22.2 (8.1) | 38 (7) | 13:45 |
| Site 2 - NT | 44 | 19.5 (6.06) | 28 (16) | 44:43 |
| Site 2 - Autism | 58 | 23.3 (8.69) | 44 (14) | 58:53 |
| Site 3 - NT | 13 | 21.7 (5.77) | 11 (2) | 13:13 |
| Site 3 - Autism | 16 | 21.7 (7.43) | 10 (6) | 16:16 |
| Site 4 - NT | 8 | 23.4 (15.1) | 7 (1) | 8:7 |
| Site 4 - Autism | 16 | 26.1 (6.72) | 11 (5) | 16:16 |
| Total - NT | 76 | 21.3 (7.78) | 51 (25) | 73:76 |
| Total - Autism | 130 | 22.7 (8.08) | 97(33) | 103:130 |

### MRS data acquisition

MRS data were acquired at four 3T MRI sites (one GE, three Siemens; Table S1) during wakefulness. A T_1_-weighted MP-RAGE anatomical image was acquired for voxel placement and tissue segmentation (TR/TE/TI = 2300/3/900ms, voxel size = 1.1 × 1.1 × 1.2 mm, flip angle = 9-11°, matrix size = 256 × 256, FOV = 270 mm, 176-208 slices). MRS data were acquired from a 26 x 40 x 24 mm thalamus voxel and a 35 x 30 x 25 mm ACC voxel, both centred on the midline (Fig 1). MRS data were acquired using vendor native PRESS; TR/TE: 2000/35ms, 64 averages, 4096 datapoints sampled at 4000Hz spectral bandwidth, followed by universal HERMES; 20 ms editing pulses at 1.90 ppm, 4.56 ppm and 7.46 ppm in the ON_GABA,_ ON_GSH_ and OFF conditions respectively, TR/TE=2000/80 ms, 240 averages (60 averages per sub-experiment), 4096 datapoints sampled at 4000 Hz spectral bandwidth (48). Water unsuppressed acquisitions were acquired (16 transients) for eddy-current and frequency correction Table S1 - (49,50). Data were acquired during wakefulness. ACC and thalamus voxel numbers acquired per site were largely consistent, excluding site 1, where a Thalamus voxels were no longer collected after year 2 (Table 1; a consequence of available scan time reductions).

**Figure 1.**
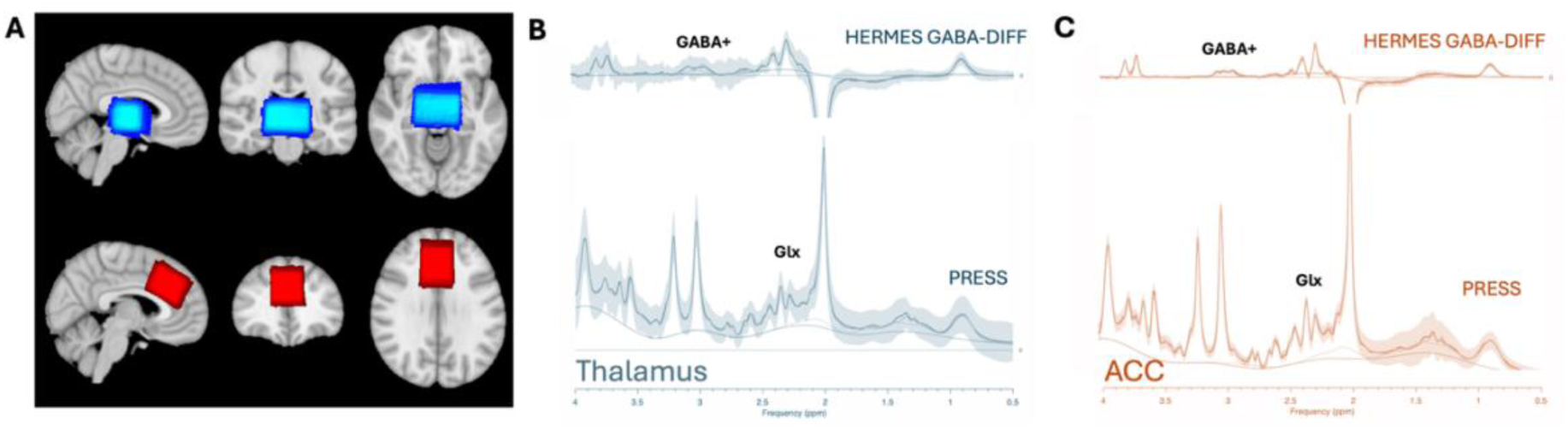
Voxel placement and mean HERMES and PRESS spectra. (**A**) Standard T_1_-weighed image (MNI52-T_1_-1 mm) with heat-plots depicting thalamus (blue) and ACC (red) MRS voxel overlap across all participants. Greater intensity indicates a greater degree of voxel overlap. Heat-plots were created by registering participant T_1_-weighted images and voxel masks to a standard space (MNI52 T_1_ 1 mm brain). (**B**) Mean HERMES GABA-DIFF and PRESS spectra showing model fit, model baseline and median residual fits (error bars) for the thalamus voxel. (**C**) Mean HERMES GABA-DIFF and PRESS spectra showing model fit, model baseline and median residual fits (error bars) for the ACC voxel. GABA+ and Glx spectral peaks are indicated. Note mean spectra per site are reported in Fig S1.

**Figure 1.**
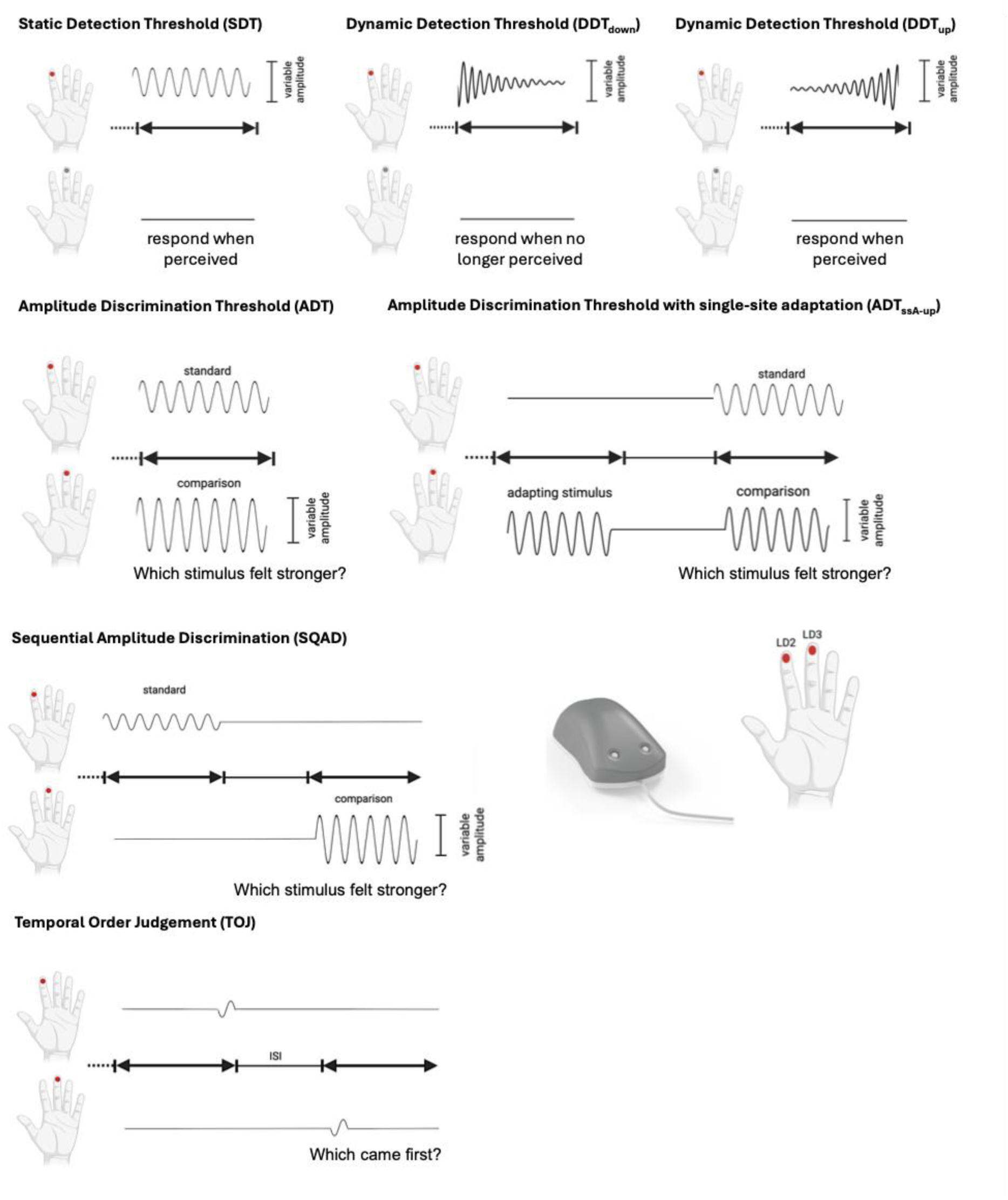
The vibrotactile psychophysics battery. Vibrotactile tasks performed using the Cortical Metric Brain Gauge Stimulator, which delivers vibrotactile stimulation to the index (LD2) and middle (LD3) finger of a participant’s left hand. Participants responded by clicking on the left or right buttons of a mouse using their right hand. ISI = interstimulus interval.

### MRS data processing

MRS data were processed using Osprey (Version 2.4.0; (51), an automated software for MRS analysis developed and implemented in Matlab (R2022a, The Mathworks Inc, Natick MA, USA). Pre-processing included coil combination, frequency and phase correction (50), eddy-current correction (52), and residual water removal (53). HERMES sub-spectra were aligned using the 2.01 ppm NAA peak before subtraction to calculate the GABA-DIFF spectra (A + B – C – D). Averaged PRESS and HERMES spectra were modelled with TE-specific simulated basis sets and a flexible spline baseline (generated in the MATLAB toolbox FID-A; (51,54). Basis sets for macromolecule and lipid contributions were integrated as TE-specific gaussian basis functions (51). All spectra were modelled between 0.5 ppm and 4 ppm with non-linear baseline correction and a knot spacing of 0.55 ppm according to the Osprey model algorithm. For a full list of fitted metabolites, please refer to the supplementary. Glx, GABA+, glutamate, and glutamine were the primary metabolites of interest; note that at 3T, glutamate is reported as Glx (glutamate + glutamine) due to significant spectral overlap, and GABA is quantified as GABA+ due to macromolecular contamination of the edited signal (55). Glx (and glutamate) were estimated from the PRESS spectra, while GABA+ was quantified from the HERMES GABA-DIFF spectra (Fig 1). MRS voxels were co-registered to subject-specific T1-weighted images using SPM12. Tissue-corrected, water-scaled metabolite concentrations were estimated accounting for grey matter, white matter, and CSF fractions and tissue-specific relaxation times (56), as recommended in consensus papers (57,58). T_1_-weighted images and voxel masks were registered to a standard MNI152 T_1_ 1 mm brain anatomical image for the creation of voxel overlap heat plots (Fig 1).

### Spectral artefacts and quality control (QC)

Spectra were visually assessed by experienced MRS data users (AT, NP) and excluded if significant artifacts due to motion and/or lipid contamination and/or spurious echoes were present. Quantitative quality metrics ; signal to noise ratio (SNR) of the total creatine peak, linewidth of the total creatine peak (full-width-at-half-maximum (FWHM)), residual fit of spectra to basis sets, and the frequency shift used for spectral alignment) were assessed per MRSinMRS guidelines (57), with outliers identified per scanner using interquartile range criteria (Tables S3–S5). SNR was included as a summary quality covariate in all analyses. Site was also included as a covariate to account for significant between-site differences in neurochemical levels.

### GABA and Glutamate polygenic scores

DNA from blood or saliva were extracted and genotyped at the Centre National de Recherche en Genomique Humaine (CNRGH, Paris) using the Illumina Infinium OmniExpress-24v1 BeadChip. Standard quality control procedures were applied, excluding participants with genotyping rates below 95%, excess heterozygosity, or sex mismatches, and removing SNPs with low call rates or Hardy–Weinberg equilibrium violations (p < 1×10−6) in PLINK v1.9. Approximately 17 million SNPs were imputed using the Michigan Imputation Server and HRC r1.1 reference panel. Population stratification was addressed by including the first four principal components as covariates. Gene-set polygenic scores (PGS) for glutamatergic and GABAergic pathways were computed in PRSice-2 using autism GWAS summary statistics (59), restricted to SNPs within curated GABA and glutamate gene sets (Table S8 & S9; (33). SNPs were clumped at r²=0.8 within a 1Mb window, and all SNPs regardless of GWAS association threshold were included to maximise polygenic signal. Each participant’s PGS was weighted by effect sizes from the base GWAS and adjusted for the four principal components.

### Vibrotactile psychophysics

Perceptual sensitivity in the tactile domain was assessed using a Cortical Metrics Brain Gauge stimulator which applied a battery of 7 vibrotactile tasks (total duration ∼ 20 mins; (20)). The stimulator was used to deliver vibrotactile stimuli within the flutter range (10 – 50 Hz) and an amplitude of 0 – 350 μm to the glabrous skin of digit 2 (LD2 – index finger) and digit 3 (LD3 – middle finger) of the left hand via two 5 mm diameter cylindrical plastic probes (Fig 2). The vibrotactile battery consisted of three conditions for detection thresholds: static detection threshold (SDT), dynamic detection threshold with an increasing subthreshold stimulus (DDT_up_), dynamic detection threshold with decreasing subthreshold stimulus (DDT_down_). There were also three conditions for amplitude discrimination: simultaneous amplitude discrimination with no adapting stimulus (ADT), simultaneous amplitude discrimination with a single-site adapting stimulus (ADT_ssA-up_), sequential amplitude discrimination (SQAD), and one condition for the temporal order judgement task (TOJ; Fig 2).

Participants responded to trials using a mouse. An adaptive stair-case tracking paradigm was used. For more details on the vibrotactile battery used please refer to (5,21,35,60).

Raw data were analysed using a custom-made software package in R (available at: https://github.com/HeJasonL/BATD). Quality of vibrotactile psychophysical measures was assessed using response tracking plots and box plots of participant thresholds, with extreme outliers removed (> ± 3 x SD; Fig S5). From the vibrotactile tasks we obtain several derived measures including estimated feed-forward inhibition engagement (the percentage difference between DDT and SDT), estimated lateral inhibition engagement (the percentage difference between ADT and SQAD), and estimated effect of adaptation (the percentage difference between ADT_ssA-up_ and ADT), described in more detail in the Supplement.

### Sensory Questionnaires

#### Sensory Experiences Questionnaire (SEQ) & Short Sensory Profile (SSP)

Sensory processing and associated behavioural responses were also assessed using the SEQ and SSP. The SEQ Version 3 (61,62) is a 105-item questionnaire (15 – 20 minutes) used to assess sensory experiences across different sensory domains. Responses are based on a 5-point Likert scale, ranging from 1 (almost never) to 5 (almost always), with higher scores indicating greater sensory difficulties. SEQ yields subscale scores (hypo-responsiveness (HYPO), hyperresponsiveness (HYPER), seeking (SEEK), sensory interests, repetitions and seeking behaviours (SIRS) and enhanced perception (EP). Note EP is characterised by superior acuity in the awareness of specific sensory stimuli (3). EPHYPER refers to a total combination score of EP and HYPER, while HYPOSIRS refers to a total combination score of HYPO and SIRS. A total score is also calculated (TOTAL). Higher scores on the SEQ indicate greater sensory challenges.

The SSP is a 34-item questionnaire which comprises 38 items, each item is scored on a 5-point Likert-rating scale from 1 (always occurs) to 5 (never occurs; (63,64). The SSP is a short form of the Sensory Profile 2 (64), and has 7 subscales which include tactile sensitivity, taste/smell sensitivity, under-responsive/seeks sensation, movement sensitivity, auditory filtering, low energy/weak, visual/auditory sensitivity. A total SSP score is also calculated. Lower scores on the SSP indicate greater sensory challenges.

## Statistical analysis

Proposed analyses were preregistered via the LEAP bottom-up proposal system before any analyses were undertaken.

Due to significant age, site and potential sex effects in neurochemical data (65,66); see supplementary materials, Table S6, we used a trajectory modelling approach to quantify how autistic individuals deviated from a predicted NT Glx/GABA+ mean while accounts for these variables. Trajectories were estimated from NT participants using a non-parametric Gaussian Process Regression (GPR) approach (67); https://github.com/ralidimitrova; see Supplement for more detail). GPR is robust to relatively small sample datasets because it models non-linear trajectories while accounting for changing uncertainty across the age range studied due to heterogeneity in sampling density. The goal of this trajectory modelling was to account for nuisance variables (age, sex, site) within our sample, and to focus on individual differences relative to an expected Glx/GABA+ trajectory, rather than group-level mean neurochemical differences. Consequently, these models were developed specifically for the current dataset and are not intended for application to independent datasets acquired on different scanners. Note that findings were largely replicated when using a more conversative linear regression approach.

Separate GPR were estimated for each of the neurochemical measures (Glx, GABA+, and Glx: GABA+ ratios) per voxel (ACC and Thalamus; see Table S7 for n per GPR). Independent variables were age, sex, and site; dependant variables were neurochemical levels. For each participant, z-scores were calculated as the difference between the observed neurochemical level and the model-predicted mean (of NT individuals; given age, sex, and site), scaled by the model-predicted standard deviation (i.e., the square root of the predicted variance). This produced z-score in standardised units, representing how far each participants neurochemical value deviated from the expected mean given the participants age at scan, site of scan and sex.

### Associations between MRS, polygenic scores, questionnaires and psychophysics

Associations between Glx, Glx: GABA+ and GABA+ age-related z-scores, polygenic scores, sensory questionnaires (SEQ and SSP) and vibrotactile thresholds were assessed using linear regressions (sensory measure ∼ z-score(*diagnosis) + SNR). Participant N number per analysis are reported in the supplementary (Table S11 & S12), as well as our sensitively analysis for this approach. Analyses were performed across groups and per group when interaction effects where significant (NT and autism). *Associations between NT neurochemical levels and NT questionnaire-based sensory reactivity scores are not reported due to more limited variation in NT sensory reactivity scores, meaning that trends were driven by single datapoints.* For polygenic scores, associations between glutamate PGS and *glutamate only* deviation scores were additionally assessed to observe if glutamine in the Glx signal masked any significant associations.

Results were corrected using False Discovery Rate (FDR) corrections were appropriate, while we assessed the stability of any interaction effects, using non-parametric bootstrap resampling (5,000 iterations) to estimate bias-corrected and accelerated (BCa) 95% confidence intervals for the interaction coefficients. If the CI excludes zero, this is evidence that the effect is consistently estimated across resamples.

### Associations between vibrotactile and sensory questionnaire measures

Spearman’s rank correlation coefficients were calculated to assess correlations between vibrotactile thresholds and SEQ and SSP questionnaire scores. Correlations between vibrotactile thresholds and SEQ/SSP scores are not reported for the NT group due to more limited variation in NT sensory reactivity scores. To assess whether these associations were independent of demographic and site-related factors, analyses were repeated using linear regression models adjusting for age, sex, and site.

## Results

We used Gaussian Process Regression (GPR) to calculate individual Glx, GABA+, and Glx:GABA+ ratio deviation Z-scores relative to the age-, sex-, and site-adjusted neurotypical (NT) model-predicted mean (see Fig S2). Neurochemical Z-scores were extracted for both autistic and NT participants and were used in all subsequent analyses. Descriptive summaries of thalamic and ACC Glx, GABA+, and Glx:GABA+ levels, including diagnostic group and age-related differences, are provided in the Supplementary Material (Fig. S2; Table S6), as characterising group differences was not the aim of the present analyses.

## Associations between Glx and GABA+ z-scores and polygenic scores

First, we investigated whether individual neurochemical z-scores were associated with common genetic variation in GABAergic and glutamatergic gene sets, summarised as polygenic scores (PGS). Note here we include GABA, Glx and *glutamate-only* z-scores. Glutamate-only Z-scores were included as a sensitivity analysis to assess whether associations between glutamatergic PGS and Glx reflected the glutamate rather than glutamine component of the composite Glx signal. However, because glutamate-specific measurements at 3T can be less reliable than composite Glx estimates, results below preferentially focus on Glx findings.

Across all participants, higher glutamate PGS was associated with more negative (or less positive) Glx z-scores across all participants (beta = −0.57, p = 0.04), however this did not survive multiple comparison correction, and no significant interaction with diagnostic group was observed. In contrast, glutamate PGS significantly interacted with diagnosis to predict thalamic *glutamate-only* z-scores (beta_interaction_ = 0.41, p_adjusted_ = 0.02; Fig S4). Post-hoc stratified analyses showed that higher glutamate PGS (reflecting increased autism liability within glutamatergic gene sets) was associated with more negative (or less positive) thalamic glutamate z-scores in NT individuals (beta = –0.35, 95% CI [−0.600, −0.102], p = 0.01), corresponding to lower glutamate levels relative to age-, sex- and site-expected values. This association was not observed in autistic individuals (beta = 0.049, 95% CI; [-0.166, 0.264], p = 0.65).

No significant associations were observed between thalamic GABA+ deviations and GABA PGS, nor between ACC neurochemical deviations and their corresponding PGS in either group.

## Associations between Glx and GABA+ z-scores and vibrotactile perceptual thresholds

Next, we investigated whether individual neurochemical z-scores were associated with vibrotactile thresholds. Across all participants, multiple diagnosis × neurochemical interactions were observed in predicting vibrotactile thresholds (Fig 3), indicating that the relationship between regional E/I neurochemistry and tactile perception differed by diagnostic group. To assess the stability of these interaction effects and reduce reliance on the assumptions of ordinary least squares regression, we performed non-parametric bootstrap resampling (5,000 iterations) and estimated bias-corrected and accelerated (BCa) 95% confidence intervals for the interaction coefficients. If the CI excludes zero, that’s evidence the effect is consistently estimated across resamples.

**Figure 3.**
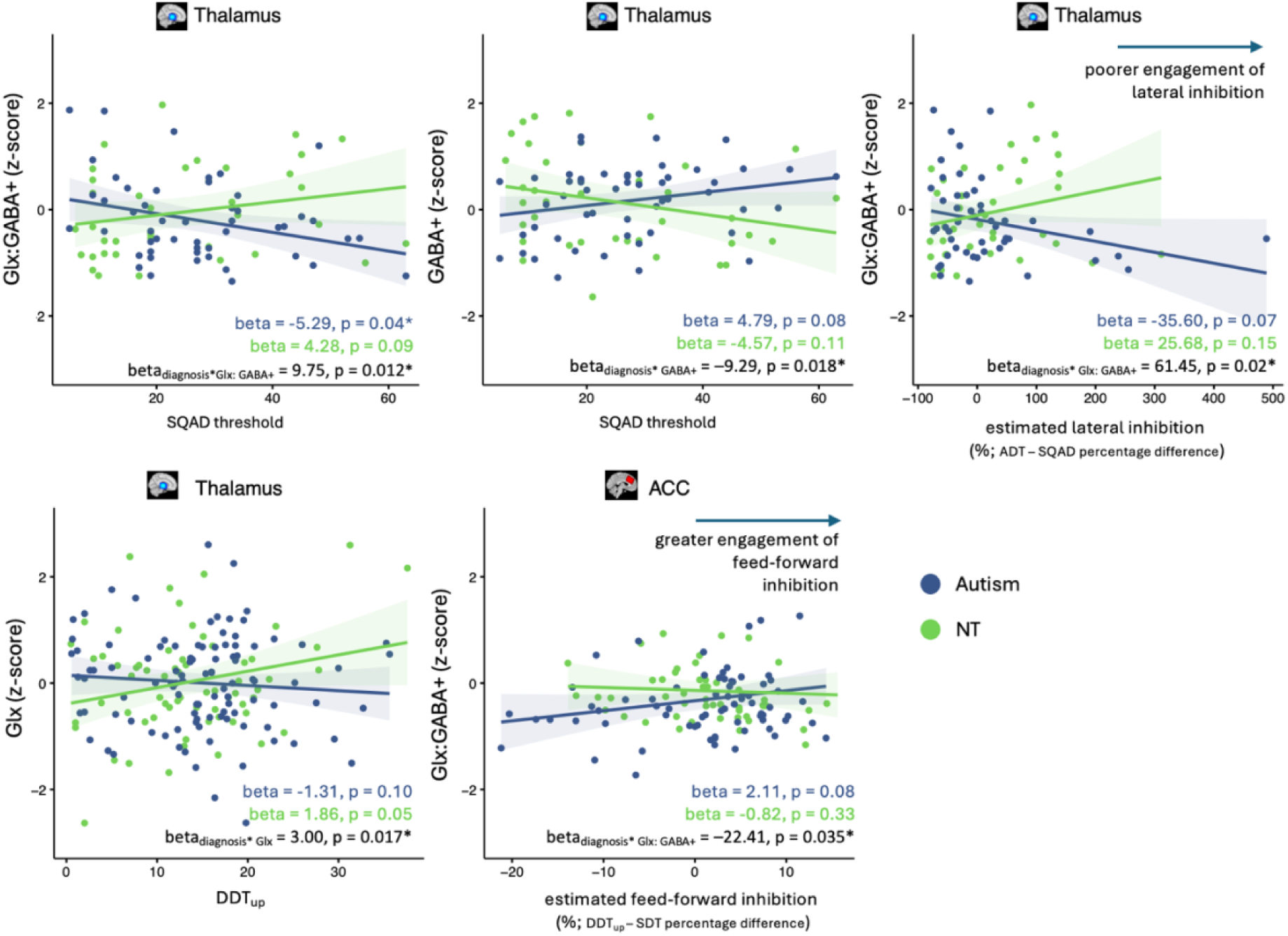
Diagnosis moderates the association between Glx, Glx:GABA+ and GABA+ z-scores and vibrotactile perceptual thresholds. Individual ACC and thalamic Glx, Glx:GABA+ and GABA+ z-scores were associated with vibrotactile thresholds. Across all participants, significant diagnosis × neurochemical interactions were observed when predicting vibrotactile thresholds, indicating that the relationships between Glx, Glx:GABA+ and GABA+ z-scores and vibrotactile thresholds differ between autistic and NT individuals. Significant interactions are shown (black text, right-hand corner), as well as corresponding beta values per diagnostic group (blue/green text, right-hand corner). All analysis controlled for quality of MRS data (SNR).

Interaction effects were most pronounced for thalamic Glx:GABA+ and GABA+ z-scores in relation to SQAD. Specifically, significant diagnosis interactions were observed for; thalamic Glx:GABA+ z-scores predicting SQAD threshold (beta_diagnosis*Glx: GABA+_ = 9.75, p = 0.012; 95% BCa CI: [1.624, 17.192]), thalamic GABA+ z-scores predicting SQAD threshold (beta_diagnosis* GABA+_ = –9.29, p = 0.018; 95% BCa CI: [-16.129, -1.709]), thalamic Glx:GABA+ z-scores predicting estimated lateral inhibition engagement (ADT–SQAD percentage difference; beta_diagnosis* Glx: GABA+_ = 61.45, p = 0.02; 95% BCa CI: [13.38, 103.27]), thalamic Glx z-scores predicting DDT_up_ (beta_diagnosis* Glx_ = 3.00, p = 0.017; 95% BCa CI: [0.28, 6.08]), and ACC Glx:GABA+ z-scores predicting estimated feedforward inhibition engagement (DDT_up_–SDT percentage difference; beta_diagnosis* Glx: GABA+_ = –22.41, p = 0.035; 95% BCa CI: [-45.46, 1.33). Stratified analyses confirmed that the direction of neurochemical–perceptual associations differed between autistic and neurotypical individuals (Fig 3). Raw vibrotactile thresholds and diagnostic group differences are reported in Fig S5 & Table S13. Bootstrap confidence intervals excluded zero for all significant interactions (bar the last), supporting the robustness of the observed effects.

## Associations between Glx and GABA+ z-scores and sensory reactivity (autism only)

Regional Glx z-scores associated with questionnaire-based sensory reactivity in autistic participants (Fig 4). More negative ACC Glx z-scores were significantly associated with greater sensory hypo-reactivity and sensory interests, repetitions and seeking behaviours on the SEQ (beta_SEQ-HYPO_ = −1.02, p_SEQ-HYPO_ = 0.047; beta = −1.81 _SEQ_-_HYPOSIRS_, p = 0.027 _SEQ-HYPOSIRS_). More negative ACC Glx z-scores were also associated with greater difficulties on the SSP low energy/weak subscale (beta = 1.80, p = 0.033). Similarly, more negative *thalamic* Glx z-scores were associated with greater difficulties on the SSP low energy/weak subscale (beta = 3.04, p = 0.01). Thus, lower glutamate levels relative to age- and sex-expected neurotypical means are associated with sensory reactivity differences within autistic individuals. No significant associations between GABA+ or Glx:GABA+ z-scores and sensory reactivity were observed.

**Figure 4.**
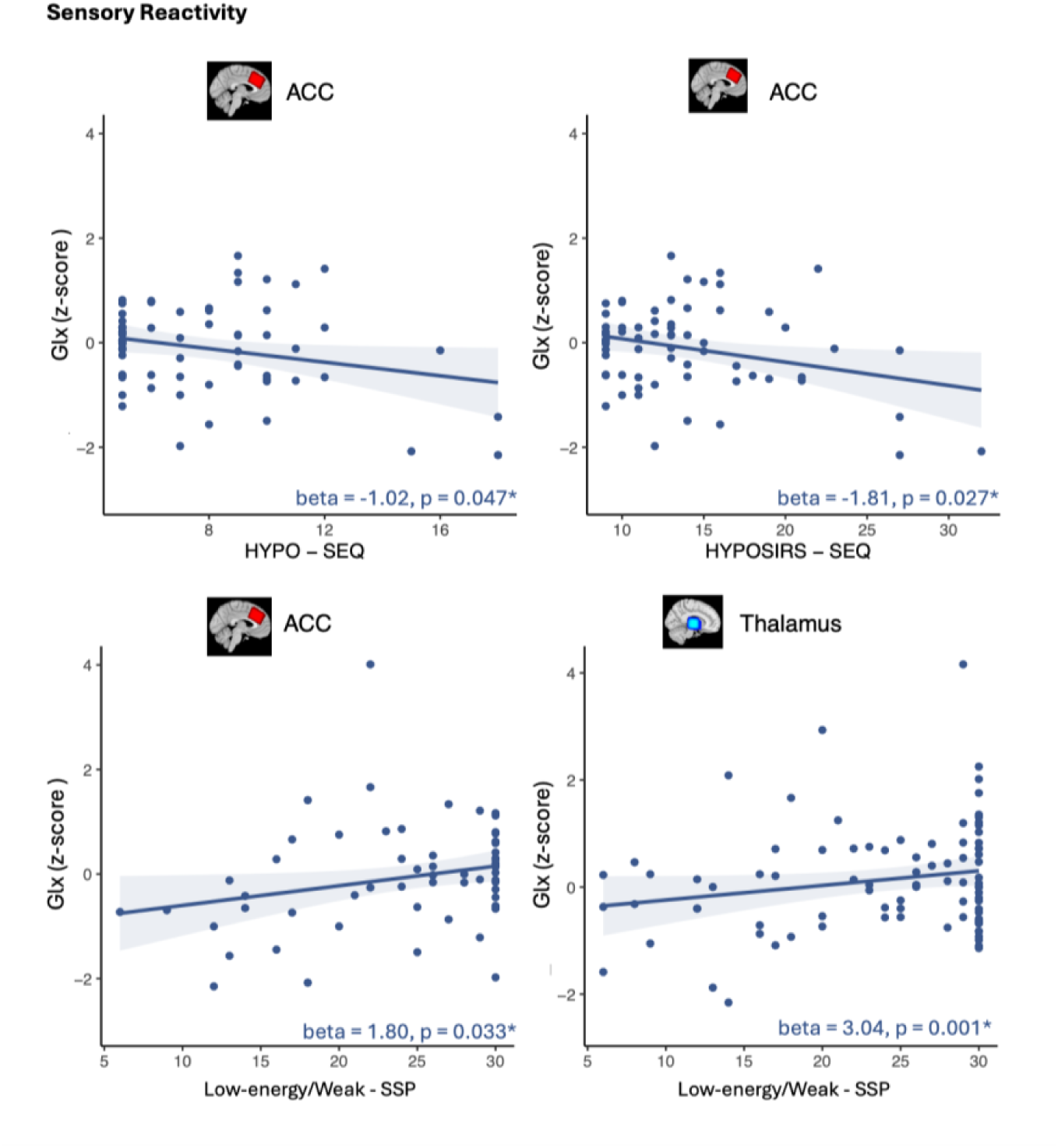
Significant associations between questionnaire-based measures of sensory reactivity and Glx z-scores. Sensory questionnaire scores (SEQ & SSP) were associated with ACC and thalamic Glx, GABA+, Glx:GABA+ z-scores in autistic participants. Only associations with Glx were significant, these are are shown (p < 0.05). Note lower scores on the SSP but higher scores on the SEQ are indicative of a greater sensory behavioural challenge.

## Associations between vibrotactile perceptual thresholds and sensory reactivity (autism only)

Spearman’s rank correlation coefficients were calculated between vibrotactile thresholds and SEQ/SSP questionnaire scores in autistic participants only (Fig 5).

**Figure 5.**
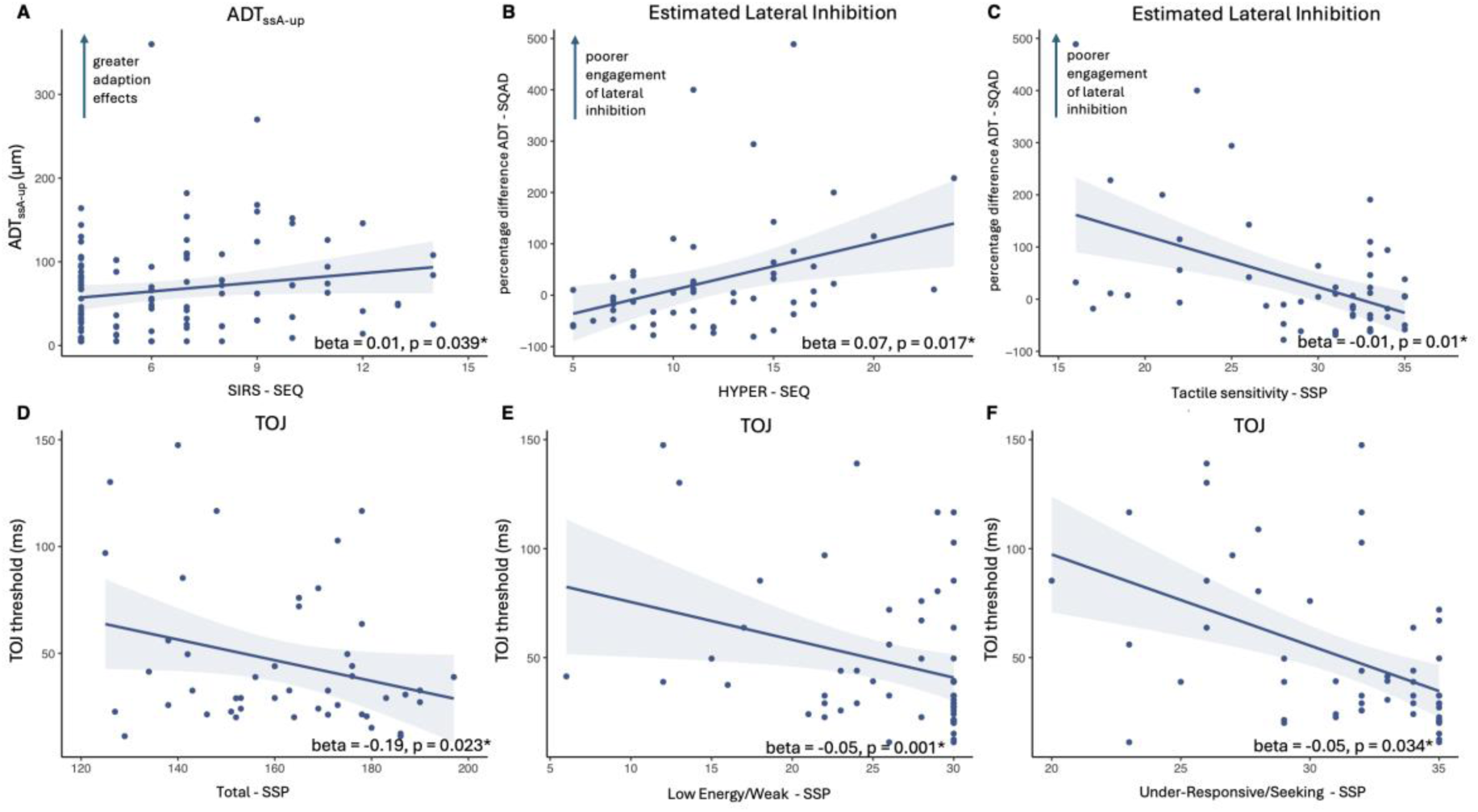
The associations between vibrotactile thresholds and questionnaire-based measures of sensory reactivity across autistic participants. Linear associations between vibrotactile thresholds and sensory reactivity scores from the SSP and SEQ, controlling for age, sex, and site. This analysis was performed across autistic participants only. Only significant findings are shown.

### For the SEQ

Higher amplitude discrimination thresholds after an adapting stimulus (ADT_ssA-up_) were associated with greater sensory hypo-reactivity, greater sensory interests, repetitions, and seeking behaviours, and greater total sensory challenge (SIRS: rho = 0.22, p = 0.021; SEQ total score: rho = 0.20, p = 0.036; HYPOSIRS: rho = 0.17, p = 0.048). Relatedly, greater effects of an adapting stimulus on amplitude discrimination thresholds (percentage difference between ADT and ADT_ssA-up_) were associated with greater total sensory challenge (SEQ total score: rho = -0.20, p = 0.044). Poorer engagement of lateral inhibition (greater percentage difference between ADT and SQAD threshold) was associated with greater sensory hyper-reactivity (HYPER: rho = 0.41, p = 0.00; EPHYPER: rho = 0.35, p = 0.01) and greater total sensory challenge (SEQ total score: rho = 0.31, p = 0.022).

After controlling for age, site and sex the association between amplitude discrimination thresholds after an adapting stimulus (ADT_ssA-up_) and SIRS (beta = 0.0097, p = 0.039), and poorer engagement of lateral inhibition (greater percentage difference between ADT and SQAD threshold) and HYPER (beta = 0.067, p = 0.017) remained significant.

### For the SSP

Higher TOJ thresholds were associated with greater overall sensory challenge (SSP total score: rho = –0.30, p = 0.028; beta = -0.19, p = 0.023 when controlling for sex, age, site)), greater SSP low Energy/Weak difficulties (rho = –0.33, p = 0.00058; beta = -0.047, p = 0.00072 when controlling for sex, age, site), and greater Under-responsiveness/Seeks Sensation difficulties (rho = –0.44, p = 0.00024; beta = -0.049, p = 0.034 when controlling for sex, age, site). Poorer engagement of lateral inhibition (greater percentage difference between ADT and SQAD threshold) was associated with greater SSP Tactile Sensitivity (rho = –0.38, p = 0.0016; beta = -0.086, p = 0.0088 - controlling for sex, age, site) and greater total sensory challenge (SSP total score; rho = –0.29, p = 0.035; NS when controlling for sex, age, site).

Thus, individual differences in vibrotactile perceptual thresholds were systematically associated with questionnaire-based measures of sensory hypo- and hyper-reactivity within the autistic group. Note however that effect sizes are minimal, and p-values are not corrected for multiple comparison correction, as tested associations are supported by previous works showing associations between tactile sensitivity and sensory reactivity (4,68–71).

## Discussion

In this study, we investigated whether regional neurochemical proxies of excitatory–inhibitory (E/I) signalling were associated with autism genetic liability indexed by glutamatergic- and GABAergic-specific polygenic scores, and with sensory phenotypes assessed using both vibrotactile psychophysical measures of tactile perception and questionnaire-based measures of sensory reactivity. Across analyses, three key findings emerged: (i) common genetic liability for autism within glutamatergic pathways predicted thalamic glutamate differences in NT, but not autistic, individuals, potentially suggesting altered genotype-neurochemistry coupling in autism; (ii) associations between regional E/I neurochemistry and tactile perception differed in direction across diagnostic groups, indicating that similar perceptual outcomes may arise from distinct circuit mechanisms shaped by altered E/I development; and (iii) perceptual differences in autistic individuals were associated with questionnaire-based measures of sensory reactivity. Together, these findings provide multi-level evidence linking genetic variation, neurochemistry, perceptual processing and sensory reactivity behaviour.

We first demonstrate that common genetical liability for autism in glutamatergic gene sets was differentially associated with thalamic glutamate z-scores between diagnostic groups. Within neurotypical participants, higher glutamatergic polygenic scores were associated with lower thalamic glutamate levels, while we failed to detect this association in autistic participants. This potential difference in coupling between regional excitatory neurochemistry and common genetic variation in autism may reflect altered genotype–phenotype relationships, potentially arising from circuit-level compensation, and/or differences in epigenetic regulation of the genome, which is influenced by environmental factors such as inflammation (72–74). Although we do not report *glutamate-only* estimates in any additional analysis in this manuscript (due to their limited reliability at 3T), our findings suggest that *glutamate-only* measures may typically better capture genetically driven differences in excitatory tone compared to Glx (which failed to associate with glutamate PGS), potentially owing to the contribution of the glutamine to the Glx signal, which may add noise to associations with glutamate PGS. This however requires replication. Similarly, macromolecular contributions to the GABA+ signal may also obscure associations between individual GABA+ z-scores and GABAergic PGS. We did not include GABA-only estimates, as macromolecule basis functions used for this correction remain approximations of *in vivo* spectra (51,75,76). Alternatively, our included sample size may have been insufficient to detect consistent associations per and across diagnostic groups (Öngür et al., 2011). Another possibility is that in both NT and autistic participants, compensatory processes beyond the level of gene expression, such as experience-dependent plasticity, modulate cortical MRS-measured neurochemistry and thereby weaken gene– neurochemical associations. Finally, while we did not account for regional gene expression, genes within the tested GABA and glutamate gene sets are likely differentially expressed in the ACC compared to the thalamus, which may bias our findings (77). Nevertheless, we are the first to probe whether genotype–*in vivo* neurochemistry coupling differs in autism, which is a step towards separating inherited from environmentally mediated contributions to excitatory-inhibitory neurochemical balance.

We next examined whether thalamic and ACC GABA+ and Glx were associated with vibrotactile perceptual performance. We found that Glx/GABA+–perception associations varied not only by diagnostic group but also by task, underscoring the importance of considering individual perceptual measures as indexing partially distinct circuit processes (consistent with prior work; (70). Tactile perception predominantly occurs in the cortex (78), where neuronal firing rates and spiking synchrony, which encode vibrotactile stimulus properties, are modulated by thalamocortical structural and functional connectivity (40,78–83). Consistent with prior reports, autistic participants showed elevated vibrotactile detection thresholds (22,70), potentially reflecting differences in how thalamocortical circuits support perception (84). Such differences align with evidence from animal models and human neuroimaging demonstrating altered thalamocortical connectivity in autism (85–93), as well as recent work in Fmr1−/y mouse models, demonstrating that tactile hyposensitivity as a result of impaired cortical sensory encoding rather than peripheral sensory deficits (94).

The difference between dynamic and static detection thresholds has been proposed as a more specific behavioural index of thalamocortical interactions, specifically feed-forward inhibition, engaged by subthreshold stimulation which preferentially recruits cortical inhibitory interneurons through strong synaptic input from first-order thalamic nuclei (21,35,70,95–97). Activated inhibitory interneurons inhibit neighbouring excitatory cells, limiting cortical excitability and increasing detection thresholds (95,97–99). Consistent with this, detection thresholds after subthreshold stimulation (dynamic detection) were significantly greater than detection thresholds without subthreshold stimulation (static detection) across all participants.

Using this proxy for thalamocortical interactions, we observed opposing relationships between regional E/I neurochemistry and tactile perception across groups. In autistic individuals, increased regional excitatory tone (a more positive Glx z-score in thalamus, and a more positive Glx:GABA+ z-score in ACC) was associated with stronger subthreshold effects in detection tasks (DDT_up,_ and (DDT_up_ – SDT)), whereas the opposite relationship was observed in neurotypical participants. As there was no difference in subthreshold effects across groups, these findings suggest that similar perceptual outcomes may be potentially supported by distinct thalamocortical circuit mechanisms, producing the observed interaction between diagnostic group, thalamic and ACC E/I balance and vibrotactile detection performance.

Such differences may reflect altered thalamocortical connectivity in autism (discussed above), which mediates sensory gating but also influences broader cortical circuit maturation (100). Consistent with this account, differences in the cortical inhibitory architecture, including alterations in GABA_A_ receptor expression (41,101–103) and interneuron density, have been widely reported in autistic individuals. Work in Fmr1−/y mouse model of syndromic autism has also shown that alterations in cortical circuit dynamics, rather than peripheral sensory deficits, can drive tactile detection differences (94). Altered E/I development itself may thus contribute to divergent thalamocortical and cortical inhibitory engagement between groups, leading to group-specific associations between regional E/I balance and tactile detection performance.

We also observed trends toward elevated tactile amplitude discrimination thresholds in autistic adolescents and adults, consistent with prior reports (4,22,70). Differences in tactile discrimination have been linked to lateral inhibition within S1, which is engaged when tactile judgments require integration across adjacent fingers (29). Post-mortem studies of autistic individuals report reduced lateral inhibitory connectivity between cortical mini-columns (41,103,104,106–109). The difference between simultaneous and sequential amplitude discrimination has previously been used as a specific *in vivo* readout of cortical lateral inhibition, which is preferentially engaged during simultaneous but not sequential amplitude discrimination tasks (22). Using this measure, while we found no group differences in estimated lateral inhibition itself, *we again observe differing associations with thalamic E/I neurochemistry between groups*. Specifically, in autistic individuals, poorer engagement of this proxy of lateral inhibition was associated with greater thalamic excitatory tone (more positive thalamic Glx:GABA+ z-scores), while in NT individuals’ poorer lateral inhibition engagement was associated with reduced thalamic excitatory tone (more negative thalamic Glx:GABA+ z-scores). These opposing relationships mirror those observed for detection tasks described above, pointing to potential differences in the development of E/I perceptual circuitry between groups which drives the differing associations between regional E/I balance and perceptual performance. However, it is important to note that our proxy for lateral inhibition and feed-forward inhibition (above) may be influenced by processes beyond cortical lateral inhibition. For instance, sequential tasks require maintaining a stimulus ‘in memory’. If autistic individuals rely less on prior information (formally referred to as having weaker Bayesian priors) this could also affect task performance, independently of lateral inhibition (the Bayesian prior hypothesis is summarised by (110).

Further evidence for potentially altered perceptual circuitry from neurochemical associations with sequential amplitude discrimination (SQAD) performance, which differentially related to thalamic E/I (Glx:GABA+ z-scores) between groups. Typically, neuronal spike timing encodes sequential stimuli (111,112) and altered thalamocortical circuitry could disrupt this process. Nevertheless, because no group differences were observed in SQAD performance itself, this ability appears intact in our participants. Thus, while autistic and NT individuals achieve comparable outcomes on sequential discrimination tasks, again they may do so via potentially distinct mechanisms. This interpretation is consistent with recent work in the Fmr1−/y mouse model using closely related tactile discrimination paradigms, which demonstrated intact behavioural categorisation despite altered cognitive contributions to perceptual decision-making (113). Together, these findings suggest that comparable tactile performance may be achieved through different neural computations in autism, with higher-order cognitive processes influencing how sensory information is used during perceptual decisions.

Note while thalamic MRS E/I proxies appear to more strongly relate to tactile perceptual thresholds, we find that ACC excitatory tone (Glx z-score) was more constantly linked to behavioural responsivity (42,81,114,115).

This suggests a multi-level model in which distinct thalamocortical and cortical mechanisms, each with their own E/I–sensory behaviour relationships, shape the heterogeneous sensory profiles observed in autism. While out of the scope of these analyses, a closer examination of the heterogeneity in sensory phenotypes specifically (e.g., subgrouping), may, in future, be important to investigate the whether the neurobiology of autism sensory differences may vary across the spectrum, and how this is relevant for treatment.

Beyond regional E/I neurochemistry–sensory behaviour associations, our findings suggest that autism-related differences in tactile perception are behaviourally relevant, relating to sensory reactivity. In autistic individuals, poorer engagement of S1 lateral inhibition (index by the difference between SQAD and ADT performance, which was differentially associated with thalamic Glx:GABA+ z-scores across groups) was associated with greater sensory hyper-reactivity. Altered lateral inhibition circuitry may heighten the perception of, and attention to, both relevant and irrelevant sensory inputs, thereby contributing to sensory hyper-reactivity (116), consistent with prior human and animal studies linking increased sensory cortices excitatory tone to sensory hypersensitivity (21,117–119). In contrast, stronger adaption effects (on amplitude discrimination tasks) were associated with sensory-seeking profiles. Enhanced adaptation potentially reduces perceptual salience and so creates a drive a need for more intense stimulation. Together, these findings raise the possibility that different perceptual mechanisms contribute to distinct dimensions of sensory reactivity in autism. Sensory hyper-reactivity may be more closely related to altered cortical lateral inhibitory processes, whereas sensory-seeking behaviours may reflect differences in sensory adaptation. This distinction could help explain why the same individual can exhibit both hyper- and hypo-reactive behaviours within the same sensory modality, with these behavioural profiles arising from separate but co-occurring perceptual mechanisms, which our findings also suggest may differ between autistic and neurotypical individuals (5,120,121). Note, however, that these associations were modest in magnitude and were not corrected for multiple comparisons because they were based on a priori hypotheses derived from previous literature.

There are several limitations of this study to be noted. First, differences in response bias, attention, or co-occurring ADHD, common in autism, may have contributed to variability in vibrotactile performance between groups (70,122). However, if such factors fully accounted for group differences, group effects would be expected across all tasks, which was not observed. Second, MRS measures of GABA and glutamate (Glx) provide indirect proxy estimates of excitatory and inhibitory activity, and cannot distinguish between cellular, synaptic and extra-synaptic pools of glutamate and GABA, as well as their metabolic turnover (123).

Furthermore, GABA+ and Glx MRS-levels reflect known and unknown sources of variability. These include inter-scanner differences, relaxation effects, B₀ inhomogeneity, macromolecular contamination, and variation in tissue water content, all of which can influence signal amplitudes. Nonetheless, our quantification pipeline followed standard best-practice recommendations (124) and we have provided clear detail throughout the manuscript on the quantification methods used to facilitate future comparisons. MRS measurements were also taken at rest, in limited brain regions, and so do not reflect task-evoked neurochemical changes in response to vibrotactile stimulation. To evidence the functional relevance of our findings, future work is necessary to simultaneously record functional changes in regional E/I across the brain during vibrotactile paradigms.

Associations between E/I-related neurochemistry and polygenic scores were corrected for multiple comparisons because no previous studies had established specific hypotheses regarding genotype– neurochemistry relationships. In contrast, associations between neurochemical measures and sensory outcomes were not corrected for multiple comparisons. These analyses were hypothesis-driven, based on prior evidence (including our own previous work) linking regional E/I neurochemistry to sensory processing and tactile perception to sensory reactivity in autism (21,22,35,36,68,69,71,119,125–127). Moreover, the sensory battery comprised multiple psychophysical and questionnaire-based measures that capture related aspects of sensory function and are therefore not statistically independent. Accordingly, these analyses should be interpreted as targeted tests of a priori hypotheses rather than as an exploratory screen of independent associations. All primary analyses were specified a priori through the LEAP bottom-up proposal system and preregistered before data analysis. To further assess the robustness of significant interaction effects, non-parametric bootstrap resampling (5,000 iterations) was used to obtain bias-corrected and accelerated (BCa) 95% confidence intervals. Given the number of statistical tests performed, however, these findings should be interpreted cautiously and require replication.

## Conclusions

Overall, these findings indicate that the relationship between regional excitatory–inhibitory (E/I) neurochemistry and tactile perception potentially differs across diagnostic groups, suggesting differential engagement of thalamocortical and cortical perceptual mechanisms in autistic and neurotypical individuals. Although we cannot exclude the possibility that additional factors contribute to these associations, and they require replication in larger datasets, the task specificity and consistent opposite-direction effects observed across groups add weight to the interpretation that underlying perceptual circuit dynamics may differ between autistic and neurotypical participants. Within this framework, sensory differences in autism may not be downstream consequences of altered emotional or behavioural reactivity but instead reflect differences in perceptual processing. Accordingly, our findings provide evidence towards studying autistic sensory processing on its own terms, rather than primarily in reference to neurotypical norms.

This reconceptualization has important implications for future autism research by shifting the emphasis toward identifying the developmental and circuit-level mechanisms that give rise to autistic sensory processing specifically. It also has translational relevance, suggesting that interventions may be more effective when they support and accommodate autistic sensory processing, rather than attempting to normalise it toward neurotypical patterns. More broadly, these findings highlight the need to move beyond the broad and often underspecified concept of “E/I imbalance” toward a more precise mechanistic framework in which E/I is considered in developmentally, regionally and mechanistically specific manner. This is motivated by our findings that suggest that rather than representing a simple shift along a neurotypical spectrum of E/I balance, autistic sensory processing may rely on different mechanisms that support behaviour in distinct ways.

## Data Availability

All data produced in the present study are available upon reasonable request to the authors.

## Acknowledgements

The results leading to this abstract have received funding from the Innovative Medicines Initiative 2 Joint Undertaking under grant agreement No 777394 for the project AIMS-2-TRIALS. This Joint Undertaking receives support from the European Union’s Horizon 2020 research and innovation programme and EFPIA and AUTISM SPEAKS, Autistica, SFARI. NP and AT are supported through the MRC Centre for Neurodevelopmental Disorders. This study has been further supported by the Horizon2020 supported programme CANDY (Grant No. 847818).

TA was supported by an MRC Senior Clinical Fellowship [MR/Y009665/1] and the Medical Research Council (MRC) Centre for Neurodevelopmental Disorders [MR/N026063/1]. This study represents independent research in part funded by the National Institute for Health and Care Research (NIHR) Maudsley Biomedical Research Centre (BRC) at South London and Maudsley NHS Foundation Trust and King’s College London.

We thank all those involved in data collection; Zuzana Suchomelova, Mee Rim Oh, Chirag Mehra, Mei Lin Law, Sanjana Gandhi, Laura Bravo Balsa, Maria Dauvermann, Yumnah Kahn, Esme Hayes, Leona Strauss, Noel Lam, Anouk Dykstra, Kim Lamers, Marije Mars, Lucas Geelen, Lotte Beckers, Anna Praat, Feline van Aagten, Sjors Reith, Sanne Kluin, Natalie Forde, Jill Naaijen, Anna Kaiser, Sarah Baumeister, Feline van Aagten, Hana Abouzahr, Lisa Axelsson, Ruth Darby, Elin Davelaar, Richard Delorme, Louise Duquesne, Anouk Dykstra, Anne Fritz, Lucas Geelen, Amy Goodwin, Linda Girke, Amelie Guiot, Johanna Harder, Sabine Hunnius, Sanne Kluin, Julia Koziel, Kim Lamers, Aline Lefebvre, Megan Leverington, Ellika Lule Gobena, Manon Krol, Marianne Mercier, Greg Pasco, Eline van Petersen, Anna Praat, Katie Puryer, Sjors Reith, Rachel Sarr, Johanna Saterborg, Ingrid Shragge, Agnes Vallberg. The funders had no role in the design of the study; in the collection, analyses, or interpretation of data; in the writing of the manuscript, or in the decision to publish the results.

## Disclaimer

Any views expressed are those of the author(s) and not necessarily those of the funders (IHI-JU2).

## Data availability

Osprey 2.4.0 is available through: (59,104)

